# Effect of caesarean birth on perinatal mortality for singleton breech presentation in spontaneous preterm labour – a target trial emulation using Scottish health record data

**DOI:** 10.1101/2025.05.25.25328318

**Authors:** Robin Alexander, Swetha Bhaskar, Amaya Azcoaga-Lorenzo, Adeniyi Francis Fagbamigbe, Clarine YC Chow, Kevin KW Kuan, Sarah J Stock, Stefan A Unger, Ben Swallow, Colin McCowan, Holger W Unger

## Abstract

**Background:** The effects of mode of birth for women in preterm breech labour could not be successfully determined in randomised trials. We aimed to explore the effect of caesarean birth on perinatal mortality for women in spontaneous-onset preterm labour with a singleton baby presenting breech through target trial emulation.

**Methods:** A target trial emulation of a parallel group randomised controlled trial using routinely collected Scottish electronic health record data was performed. Participants were pregnant women at 24- 36 gestational weeks with a singleton breech baby, no prior caesarean birth, in spontaneous labour with a live baby at labour onset (1 January 1997 to 31 December 2019). We compared caesarean birth (intervention) to vaginal breech birth (control) in a per-protocol analysis (actual mode of birth). The primary outcome was extended perinatal mortality (intrapartum stillbirths and neonatal deaths). A multiple logistic regression model with inverse probability weight was used to adjust for measured confounders.

There were 2,092 caesarean births and 967 vaginal breech births. In the emulated trial, caesarean birth reduced extended perinatal mortality compared to vaginal breech birth (odds ratio [OR] 0.31, 95% confidence interval [CI] 0.25 to 0.39). At 24 weeks’ gestation, caesarean birth decreased the odds of perinatal death by 47.7% (OR: 0.53, 95% CI: 0.35 to 0.78). At 36 gestational weeks it was associated with an 82.1% reduction in the odds of perinatal death (OR: 0.18, 95% CI: 0.10 to 0.32). As the risk of perinatal mortality is inversely correlated with gestational age at birth, seven and 88 caesarean births were needed to prevent one perinatal death at 24 weeks and 36 weeks’ gestation, respectively.

**Conclusions:** Caesarean birth reduces the risk of extended perinatal mortality in spontaneous preterm singleton breech labour in a per-protocol trial emulation. Observational data that accurately captures planned mode of birth is required to emulate an intention-to-treat analysis.

## Introduction

At term (37 completed gestational weeks onwards), approximately 3-4% of babies present breech.^1^ Breech presentation is more common before 37 weeks, with approximately 25% and 7% of singleton pregnancies presenting breech at 28 weeks and 32 gestational weeks, respectively.^1^ Breech presentations include complete breech (both fetal hips and legs are flexed), frank breech (fetal hip is flexed, legs are extended), and footling breech (one or both legs presenting). External cephalic version to revert breech presentation to cephalic presentation is offered from 36-37 gestational weeks onwards only.^2^

There is ongoing uncertainty regarding the optimal mode of birth for women in spontaneous preterm labour with a singleton baby that is presenting breech (‘preterm breech singletons’).^3 4^ Randomised trials comparing caesarean birth (CB) versus vaginal breech birth (VBB), conducted more than two decades ago, closed due to low recruitment before reaching sample size targets.^4–7^ Key challenges that affected recruitment to these trials were clinician or patient preferences for one mode of birth over another that inhibited randomisation, and challenges relating to obtaining consent in labour, amongst others.^8 9^ The design of clinical trials to determine the optimal mode of birth for preterm breech singletons is further challenged by the presence of maternal or fetal factors that may affect equipoise, such as concurrent fetal growth restriction, hypertensive disorder of pregnancy, and diabetes.^10^ Current guidance for parents and clinicians is thus limited to cohort studies, which are highly heterogenous in terms of inclusion criteria, availability of personnel skilled and confident to facilitate VBB, and gestational age.^11–25^ By design, these studies cannot demonstrate causality. Most but not all these studies reported higher rates of adverse fetal/neonatal outcomes following VBB, and higher rates of adverse maternal outcomes (short and long-term) after CB. For women in late preterm labour, decision-making regarding mode of birth is likely influenced by findings of the Term Breech Trial.^3 26^ The Term Breech Trial reported lower rates of perinatal mortality, neonatal mortality and serious neonatal morbidity amongst women randomised to an intended CB compared to intended VBB.^26^ Conversely, for women in extremely preterm labour, clinicians may be more likely to favour VBB.^3^ The lack of evidence regarding optimal mode of birth for women in spontaneous-onset preterm labour with a singleton breech baby highlights the need the further research in this area.^27 28^

Sufficient parental acceptance and clinical equipoise that could support randomisation to VBB or CB may exist for a subset of women, provided clinical staff are trained to manage VBB.^3 8^

^28^ Until results of carefully curated clinical trials may become available, the target trial approach, which uses observational data to emulate a hypothetical randomised trial, could provide key insights into impacts of CB on perinatal outcomes and maternal outcomes.^29 30^ Trial emulation seeks to alleviate the lack of randomisation in observational studies, which lead to imbalance in maternal, clinical, and birth characteristics that may confound a causal relationship between mode of birth and birth outcomes. In outlining an ideal target trial, it becomes more clear which confounding characteristics need to be balanced across mode of birth groups to simulate randomised assignment of mothers to birth modes. Once balanced, analyses can estimate the causal relationship between mode of birth and certain birth outcomes. Of particular interest may be the impact of CB on the risks of stillbirth or neonatal death in spontaneous preterm breech labour. Unlike iatrogenic preterm birth arising from an emerging clinical need to deliver preterm to manage a fetal, maternal or obstetric condition, a substantial proportion of women in spontaneous-onset preterm breech labour may have no or only soft indications to favour one mode of birth over another at the time of presentation to hospital. This may be most prevalent at earlier gestational age and poses a substantial challenge for obstetricians.^31^ Our primary objective was to determine the effect of CB on perinatal mortality for women with a singleton breech baby presenting in spontaneous-onset preterm labour through target trial emulation.

## Materials and Methods

### Trial design, data source and linkage

We used Scottish routinely collected clinical data to emulate a target trial of CB (intervention) versus VBB (control) in women with a singleton breech baby in spontaneous preterm labour for the prevention of perinatal death. We outlined a target trial, whereby mode of birth would be “randomly assigned” and used causal inference methods to adjust for bias and imbalance inherent in the non-randomised observational data (Supplemental Table 1). ^29 32 33^ This target trial was reported in line with the CONSORT-ROUTINE checklist, a CONSORT extension for the reporting of randomised controlled trials conducted using cohorts and routinely collected data (Supplemental Table 2).^34^

Clinical data was made available through the electronic Data Research and Innovation Service (eDRIS) of the National Health Service (NHS) Scotland (Supplemental Table 3). eDRIS curates and links sources of routinely collected data from within the NHS and other government bodies. Data used within this study included the National Records of Scotland (NRS) statutory live birth register and the NRS statutory stillbirth register. Perinatal outcomes were obtained from The General Register Office (GRO) Scottish Stillbirth and Infant Death Survey (SSBID), the Mothers and Babies: Reducing Risk through Audits and Confidential Enquiries (MBRRACE) database, Scottish Birth Records (SBR database), Scottish Morbidity Records (SMR) for Neonates (SMR 11 database); maternal health data from general acute hospital discharge records (SMR 01 database); and the Maternity Inpatient and Day Case - Scottish Morbidity Record (SMR 02 database). SMRs cover diagnoses and demographic data for all patient stays and day cases in hospital for all residents in Scotland and have been validated for use in health research.^35^

Maternal records were matched using the anonymous community health index (CHI) number, a person-level identifier used across Scotland. Infant records were then matched to the mothers using existing mother/child linkage records held by eDRIS or either date of birth or date of death. The data was subsequently used to identify eligible births, mode of birth, key fetal, obstetrics and maternal characteristics, and adverse outcomes. De-identified data provided by eDRIS for the purpose of this study were accessed during the period 1 January 2021 until 31 December 2024.

### Participants

The study population consisted of pregnant women at 24-36 gestational weeks with a singleton baby presenting breech and no prior CB who presented at Scottish hospitals from 1 January 1997 to 31 December 2019 in spontaneous preterm labour with a live baby at labour onset. We first identified all singleton preterm breech births in Scotland during the period 1 January 1997 and 31 December 2019, from 24^+0^ until 36^+6^ weeks’ gestation, following spontaneous preterm labour with or without premature prelabour rupture of membranes (PPROM). Singleton pregnancies were identified from SMR 02 records which recorded the number of fetuses within the pregnancy. Breech presentation was identified using the presentation at birth recorded in the maternal discharge data. Births were identified using a combination of maternal identifier, date of birth and condition on discharge. Information was coalesced into one record when multiple records for a unique pregnancy were identified (e.g., because of patient transfers). Breech births that occurred following spontaneous onset of labour were then identified. We subsequently excluded pre-labour (antepartum) stillbirths as these cannot be attributed to mode of birth. Lastly, we excluded women with a history of one or more previous CB as in our cohort all subsequent births were by repeat CB, precluding their inclusion in the target trial.

### Interventions

The intervention tested was CB, which was compared to the control, VBB. Participants were categorised as CB or VBB according to the recorded actual mode of birth from SMR02 records. Databases did not capture information on intended/planned mode of birth.

### Outcomes

The predefined outcome for the target trial was *extended perinatal mortality,* defined as a composite of intrapartum stillbirth and neonatal death (infant death in first 28 days of life).

Fetuses who died intrapartum and neonates who died within 28 days of birth were identified using the SSBID, MBRRACE, and NRS datasets. In the absence of stillbirth or death record we identified that the baby survived beyond 28 days (Supplemental File 1).

### Statistical analysis

Based on the published literature and the subject matter expertise of the authors, we developed a directed acyclic graph (Figure 1) to conceptualise prognostic factors for mode of birth (exposure) and for extended perinatal mortality (outcome) as reported in the literature.^12 13 17 31^ Factors that occur or exist prior to birth and affect both mode of birth and extended perinatal mortality were deemed potential confounders of the relationship between exposure and outcome.

**Figure 1.**
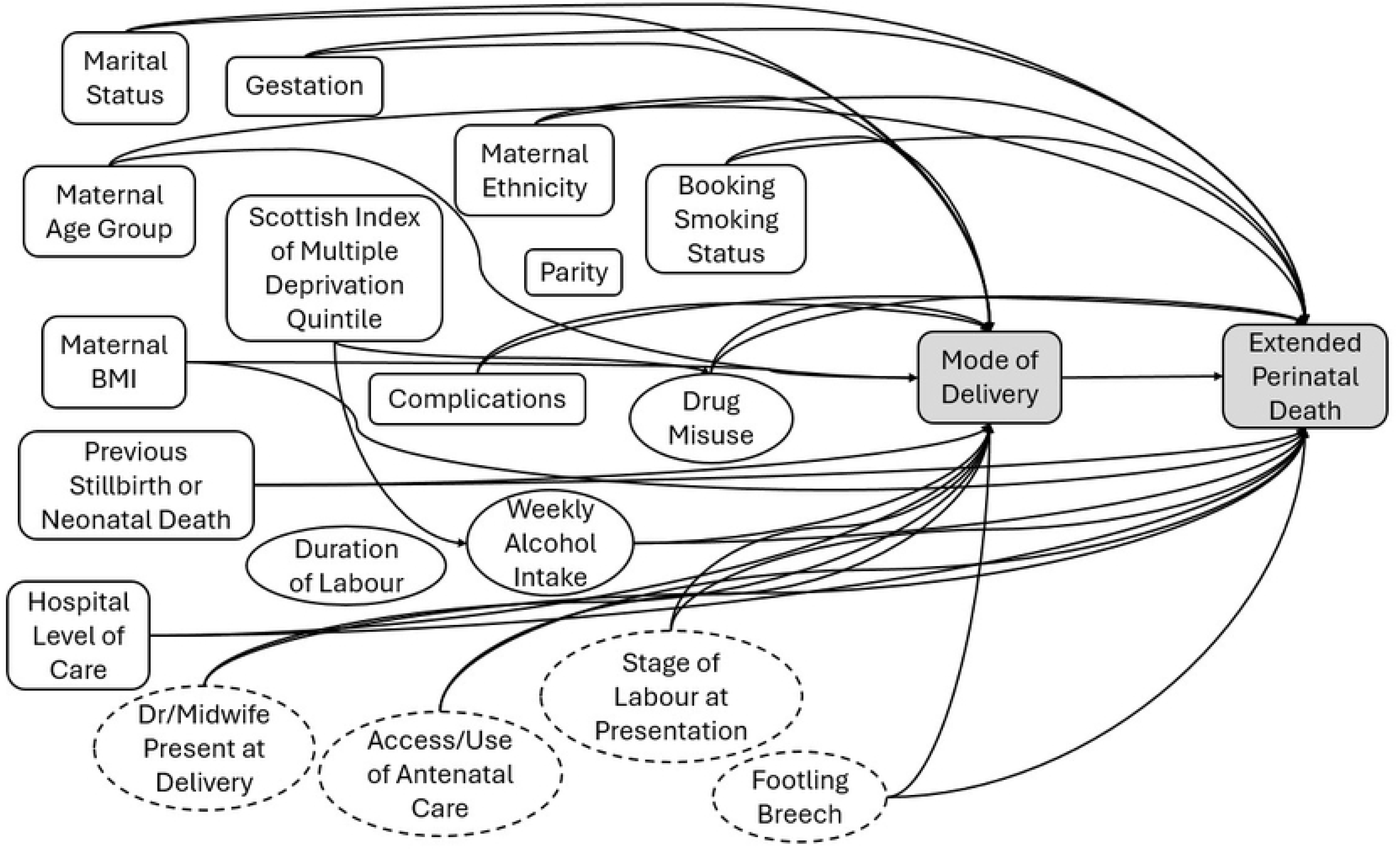
Directed acyclic graph conceptualising prognostic factors for mode of birth (exposure) and for modified extended perinatal mortality (outcome). Factors that affect both exposure and outcome confound the relationship between exposure and outcome and when measured where considered in weighted analyses. Dotted lines represent potential confounding factors that are either unmeasured or poorly documented in observational data and therefore not included in weighted analyses. (BMI, body mass index; Dr, Doctor)

Most of the identified confounding factors were available in the linked data (Figure 1). Some potential confounders, such as stage of labour at the time of assessment by clinical staff, and type of breech (e.g., frank, footling) were not documented. For the potential confounders, type of health worker present at time of birth (midwife, junior doctor, specialist) and frequency and timing of antenatal care attendance a large proportion of records had missing data, precluding their inclusion in the analysis.

Confounders considered in the analysis included maternal age, socioeconomic status (defined using quintiles of the Scottish Index of Multiple Deprivation [(SMID])^36^, maternal ethnicity, marital status, maternal body mass index at booking, smoking status at booking, parity, history of previous stillbirth and/or neonatal death, gestational age at birth (weeks), PPROM, and presence of complications. We selected complications likely known to the care provider and parents prior to birth. These included pre-existing and gestational hypertension, pre-eclampsia, pre-existing and gestational diabetes, liver disorders, large-for-gestational age fetus, fetal abnormalities, intrauterine growth restriction, infection, chorioamnionitis, placenta accreta, placenta praevia, placental abruption, antepartum haemorrhage, and rupture of the uterus were identified using the International Classification of Diseases, Tenth Revision [ICD-10] codes (Supplemental Table 4). Given the low event rates of each (Supplemental table 5), complications were aggregated into a single indicator to describe presence of any complication. This aggregate indicator was included as a potential confounding variable in the analysis. Year of birth was also adjusted for, serving as a proxy for changes in birth standard procedures and/or policies over the course of the study period. Additionally, the level of neonatal care available at the birth hospital (none, local neonatal unit, special care unit, neonatal intensive care unit) was included as a potential confounding factor.^37^ Infant sex was considered for inclusion in the analysis; however, the sex of the baby is not always known prior to birth and was thus omitted as a confounding factor. Maternal age, body mass index, and parity were discretized into clinically relevant groupings. A count of weeks preterm was used in lieu of gestational age, where 0 weeks preterm corresponded to 36 weeks’ gestation and 12 weeks preterm corresponded to 24 weeks gestation. The year of birth was centred and scaled, by subtracting the mean and dividing by the standard deviation. Correlation between the identified confounders was evaluated using Cramer’s V.^38^

A mixed effects logistic regression model, allowing random intercepts for birth hospital, was used to estimate the probability of undergoing birth by CB associated with each covariate. These probabilities were converted into inverse probability of treatment weights, which were then used to balance confounding factors across treatment groups and allow for the estimation of an average effect of mode of birth on the outcome, that is modified extended perinatal death. Weights were truncated at the 0.5^th^ and 99.5^th^ percentile to alleviate impact from large weights resulting from higher levels of imbalance in treatment across certain combined levels of confounding factors.^39^ Balance was evaluated using standardised mean differences, with standardised mean difference between-0.1 and 0.1 indicating sufficient balance.^40^ As gestational age was hypothesised to modify the effect of mode of birth on perinatal mortality,^12^ gestational age (week) was included as an effect modifier in the final model.

A final mixed effects logistic regression model adjusting for baseline confounders using inverse probability weights, allowing random intercepts for birth hospital, and including gestational age as effect modifier, was used to calculate adjusted odds ratios for the effect of CB on extended perinatal death. Heteroskedasticity and autocorrelation consistent standard errors were used in statistical tests and confidence intervals of model estimates. The number needed to treat was estimated using the adjusted odds ratio for perinatal death and unadjusted proportion of perinatal deaths in each mode of birth group.^41^

The array approach for quantifying the impact of residual confounding was conducted as a sensitivity analysis.^42^ The approach was applied using an array of all plausible prevalences of the confounder in each mode of birth group and incorporated strengths of the relationship between the unmeasured confounder and extended perinatal death that spanned from a strong protective relationship (OR 0.1) to a strong risk indicator relationship (OR 10). All analyses are compared to significance level of α<0.05. All analyses were conducted in Stata 16.1 (Stata Corp, College Station, US) and R version 4.4.3 (R Foundation for Statistical Computing, Vienna, Austria).

### Ethical approval

Ethical approval for usage of anonymised routinely collected data to enable this trial emulation was granted by the National Health Service Scotland Public Benefit and Privacy Panel for Health and Social Care (1819-0119). Ethical approval included a waiver of informed written consent for individuals included within the study. Access to the datasets by the authorised researchers was exclusively within a Safe Haven environment after de-identification of the records.

### Patient and public involvement

We had commenced the establishment of a patient advisory group to inform the design of the target trial but did not proceed due to the COVID-19 pandemic. Our study was informed by a large parent and clinician consultation study regarding mode of birth for preterm babies, including preterm breech babies, in Scotland, and reporting has been informed by recommendations on preferred language around birth.^3 43^ No parents were directly involved in setting the research question or the outcome measures, design, or implementation of the study. No patients were asked to advise on interpretation or writing up of results.

## Results

Over the study period, we identified 7,523 women who had 7,719 singleton preterm breech births (Supplemental Table 5). After excluding those who did not meet inclusion criteria (4,660 births, 4,497 women), 3,059 births (3,026 women) were included in the target trial emulation (Figure 2). There were a total of 2,092 CBs and 967 VBBs. Trial participants were largely between 25 and 34 years old (54.3%), in the lowest two socioeconomic quintiles (SIMD) (53.0%), and had no prior pregnancies (53.0%). 64.7% of breech deliveries occurred between 32 and 36 completed gestational weeks (Table 1).

**Figure 2.**
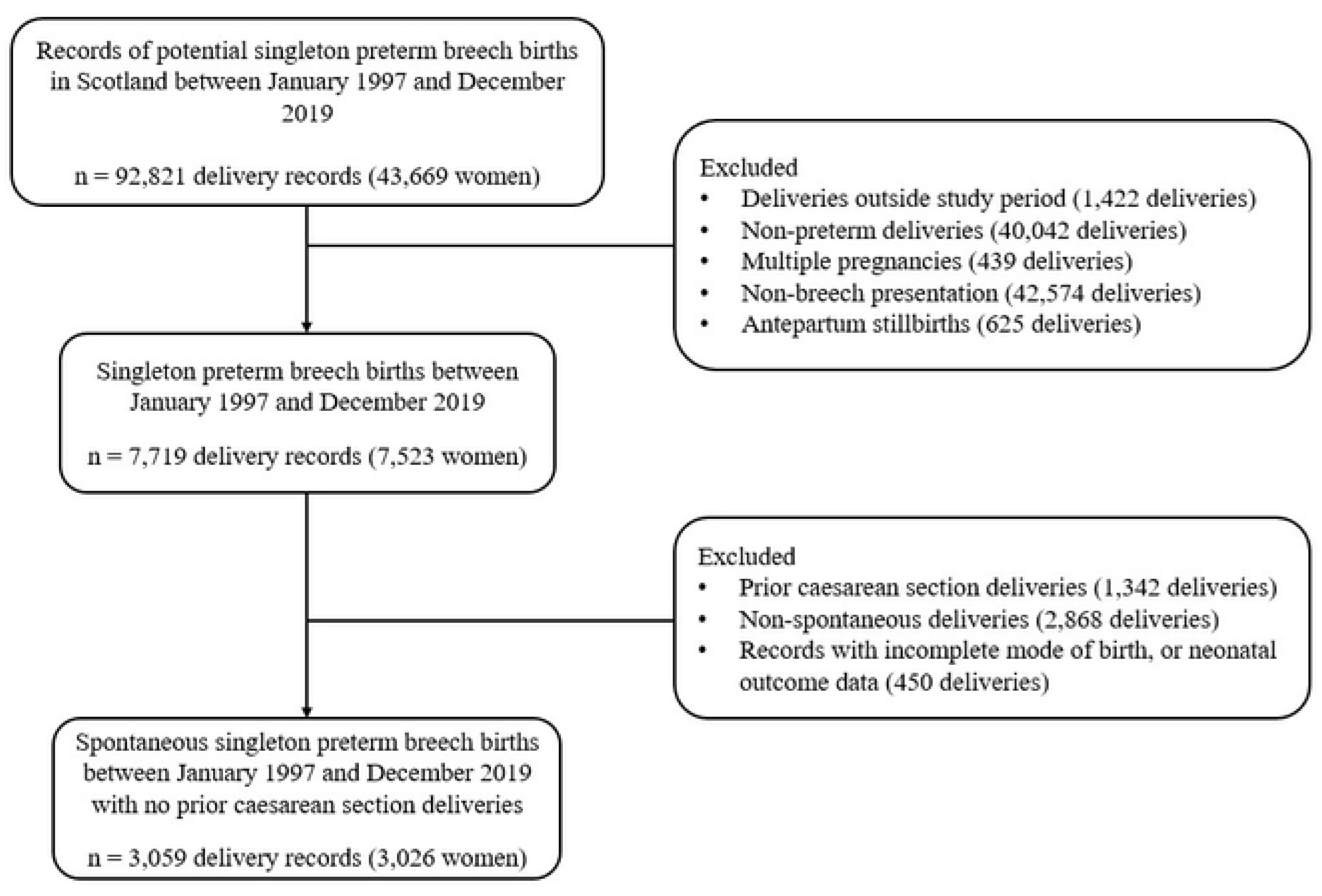
Participant selection flowchart

**Table 1.**
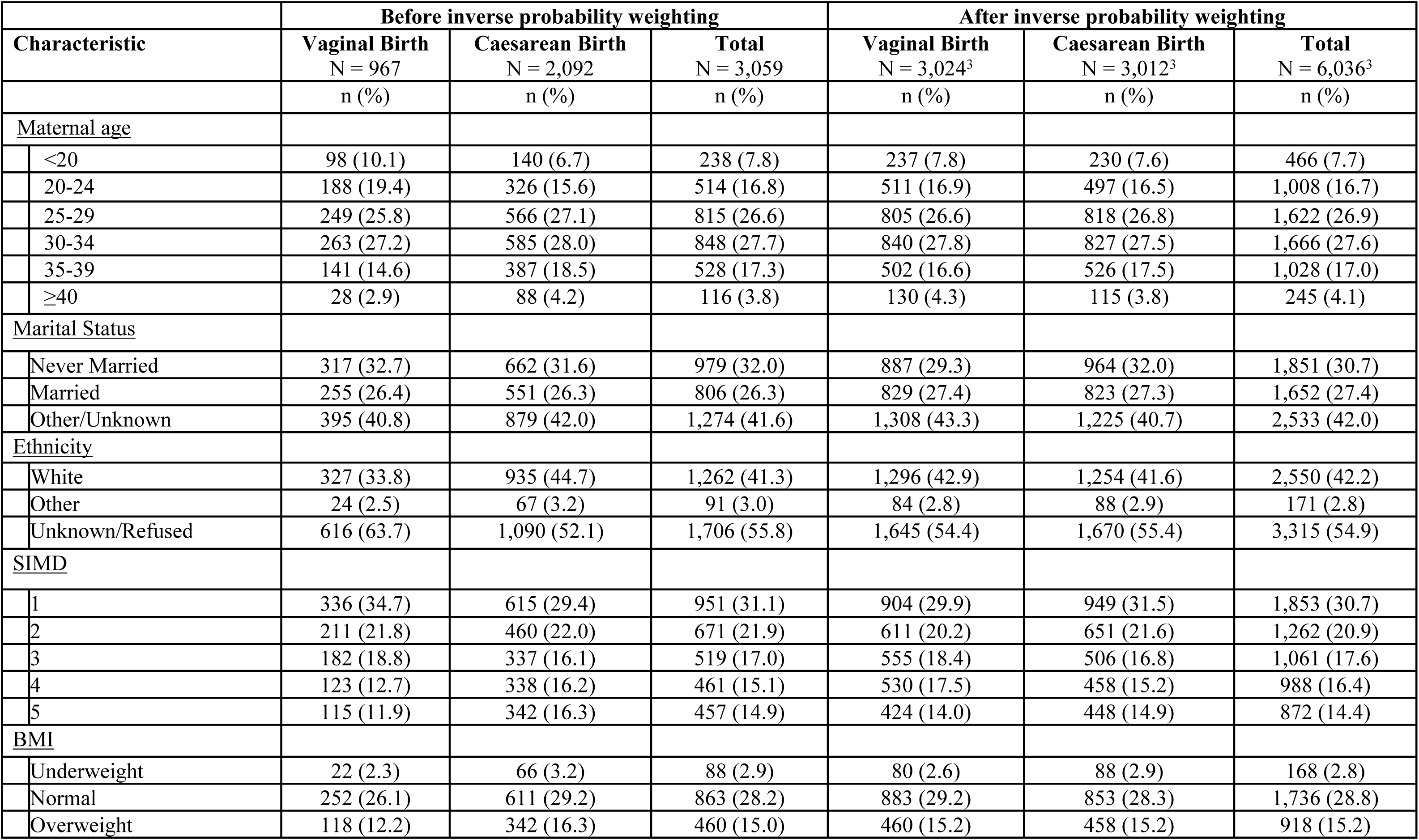
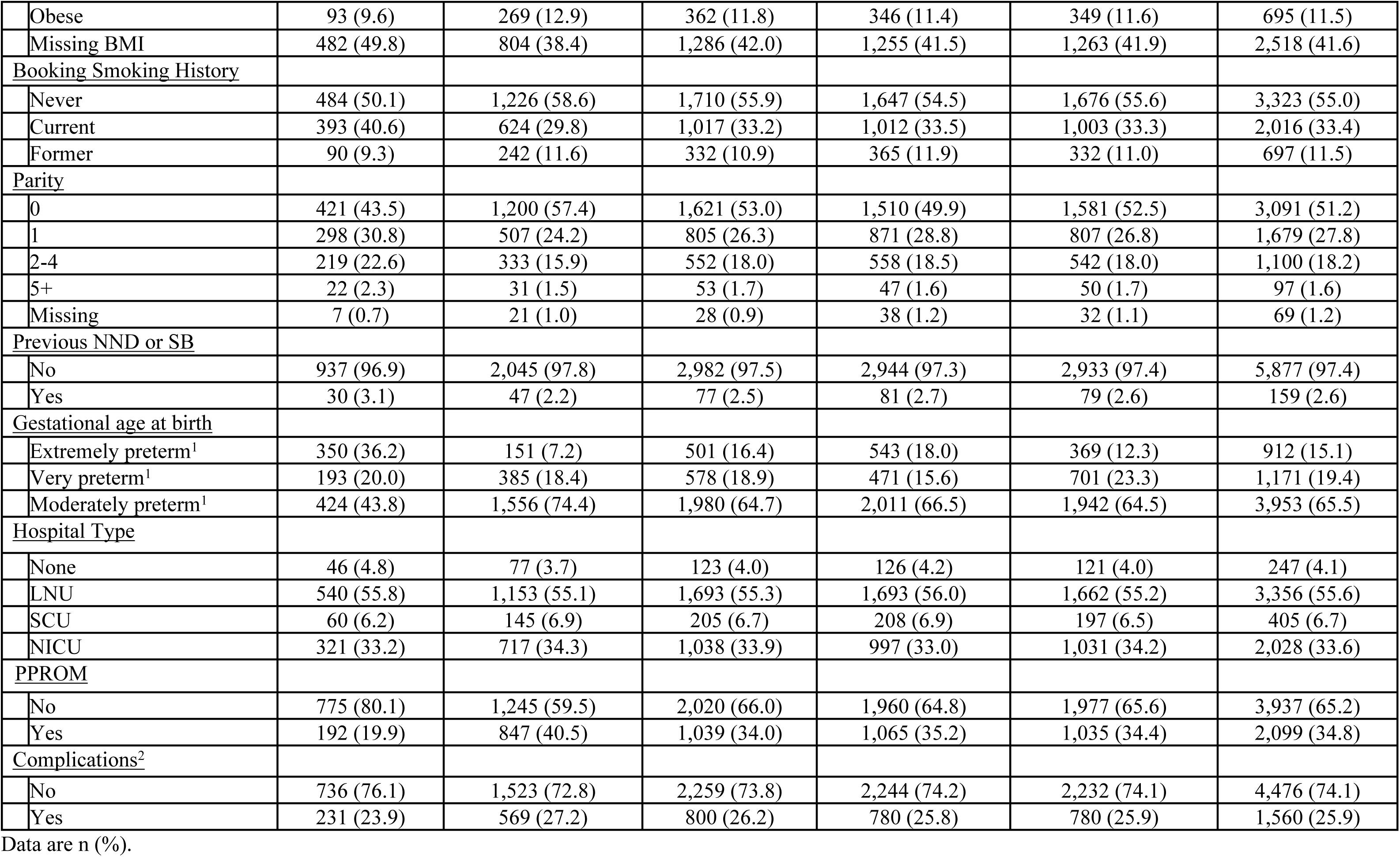
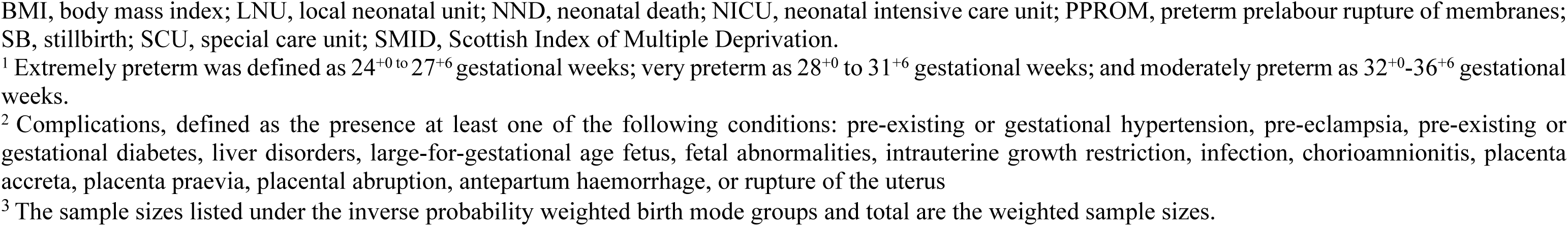
Baseline characteristics before and after applying inverse probability weighting, by mode of birth.

| Characteristic | Before inverse probability weighting |  |  | After inverse probability weighting |  |  |
| --- | --- | --- | --- | --- | --- | --- |
|  | Vaginal Birth<br>N = 967 | Caesarean Birth<br>N = 2,092 | Total<br>N = 3,059 | Vaginal Birth<br>N = 3,024<sup>3</sup> | Caesarean Birth<br>N = 3,012<sup>3</sup> | Total<br>N = 6,036<sup>3</sup> |
|  | n (%) | n (%) | n (%) | n (%) | n (%) | n (%) |
| <u>Maternal age</u> |  |  |  |  |  |  |
| <20 | 98 (10.1) | 140 (6.7) | 238 (7.8) | 237 (7.8) | 230 (7.6) | 466 (7.7) |
| 20-24 | 188 (19.4) | 326 (15.6) | 514 (16.8) | 511 (16.9) | 497 (16.5) | 1,008 (16.7) |
| 25-29 | 249 (25.8) | 566 (27.1) | 815 (26.6) | 805 (26.6) | 818 (26.8) | 1,622 (26.9) |
| 30-34 | 263 (27.2) | 585 (28.0) | 848 (27.7) | 840 (27.8) | 827 (27.5) | 1,666 (27.6) |
| 35-39 | 141 (14.6) | 387 (18.5) | 528 (17.3) | 502 (16.6) | 526 (17.5) | 1,028 (17.0) |
| ≥40 | 28 (2.9) | 88 (4.2) | 116 (3.8) | 130 (4.3) | 115 (3.8) | 245 (4.1) |
| <u>Marital Status</u> |  |  |  |  |  |  |
| Never Married | 317 (32.7) | 662 (31.6) | 979 (32.0) | 887 (29.3) | 964 (32.0) | 1,851 (30.7) |
| Married | 255 (26.4) | 551 (26.3) | 806 (26.3) | 829 (27.4) | 823 (27.3) | 1,652 (27.4) |
| Other/Unknown | 395 (40.8) | 879 (42.0) | 1,274 (41.6) | 1,308 (43.3) | 1,225 (40.7) | 2,533 (42.0) |
| <u>Ethnicity</u> |  |  |  |  |  |  |
| White | 327 (33.8) | 935 (44.7) | 1,262 (41.3) | 1,296 (42.9) | 1,254 (41.6) | 2,550 (42.2) |
| Other | 24 (2.5) | 67 (3.2) | 91 (3.0) | 84 (2.8) | 88 (2.9) | 171 (2.8) |
| Unknown/Refused | 616 (63.7) | 1,090 (52.1) | 1,706 (55.8) | 1,645 (54.4) | 1,670 (55.4) | 3,315 (54.9) |
| <u>SIMD</u> |  |  |  |  |  |  |
| 1 | 336 (34.7) | 615 (29.4) | 951 (31.1) | 904 (29.9) | 949 (31.5) | 1,853 (30.7) |
| 2 | 211 (21.8) | 460 (22.0) | 671 (21.9) | 611 (20.2) | 651 (21.6) | 1,262 (20.9) |
| 3 | 182 (18.8) | 337 (16.1) | 519 (17.0) | 555 (18.4) | 506 (16.8) | 1,061 (17.6) |
| 4 | 123 (12.7) | 338 (16.2) | 461 (15.1) | 530 (17.5) | 458 (15.2) | 988 (16.4) |
| 5 | 115 (11.9) | 342 (16.3) | 457 (14.9) | 424 (14.0) | 448 (14.9) | 872 (14.4) |
| <u>BMI</u> |  |  |  |  |  |  |
| Underweight | 22 (2.3) | 66 (3.2) | 88 (2.9) | 80 (2.6) | 88 (2.9) | 168 (2.8) |
| Normal | 252 (26.1) | 611 (29.2) | 863 (28.2) | 883 (29.2) | 853 (28.3) | 1,736 (28.8) |
| Overweight | 118 (12.2) | 342 (16.3) | 460 (15.0) | 460 (15.2) | 458 (15.2) | 918 (15.2) |
| Obese | 93 (9.6) | 269 (12.9) | 362 (11.8) | 346 (11.4) | 349 (11.6) | 695 (11.5) |
| Missing BMI | 482 (49.8) | 804 (38.4) | 1,286 (42.0) | 1,255 (41.5) | 1,263 (41.9) | 2,518 (41.6) |
| <u>Booking Smoking History</u> |  |  |  |  |  |  |
| Never | 484 (50.1) | 1,226 (58.6) | 1,710 (55.9) | 1,647 (54.5) | 1,676 (55.6) | 3,323 (55.0) |
| Current | 393 (40.6) | 624 (29.8) | 1,017 (33.2) | 1,012 (33.5) | 1,003 (33.3) | 2,016 (33.4) |
| Former | 90 (9.3) | 242 (11.6) | 332 (10.9) | 365 (11.9) | 332 (11.0) | 697 (11.5) |
| <u>Parity</u> |  |  |  |  |  |  |
| 0 | 421 (43.5) | 1,200 (57.4) | 1,621 (53.0) | 1,510 (49.9) | 1,581 (52.5) | 3,091 (51.2) |
| 1 | 298 (30.8) | 507 (24.2) | 805 (26.3) | 871 (28.8) | 807 (26.8) | 1,679 (27.8) |
| 2-4 | 219 (22.6) | 333 (15.9) | 552 (18.0) | 558 (18.5) | 542 (18.0) | 1,100 (18.2) |
| 5+ | 22 (2.3) | 31 (1.5) | 53 (1.7) | 47 (1.6) | 50 (1.7) | 97 (1.6) |
| Missing | 7 (0.7) | 21 (1.0) | 28 (0.9) | 38 (1.2) | 32 (1.1) | 69 (1.2) |
| <u>Previous NND or SB</u> |  |  |  |  |  |  |
| No | 937 (96.9) | 2,045 (97.8) | 2,982 (97.5) | 2,944 (97.3) | 2,933 (97.4) | 5,877 (97.4) |
| Yes | 30 (3.1) | 47 (2.2) | 77 (2.5) | 81 (2.7) | 79 (2.6) | 159 (2.6) |
| <u>Gestational age at birth</u> |  |  |  |  |  |  |
| Extremely preterm<sup>1</sup> | 350 (36.2) | 151 (7.2) | 501 (16.4) | 543 (18.0) | 369 (12.3) | 912 (15.1) |
| Very preterm<sup>1</sup> | 193 (20.0) | 385 (18.4) | 578 (18.9) | 471 (15.6) | 701 (23.3) | 1,171 (19.4) |
| Moderately preterm<sup>1</sup> | 424 (43.8) | 1,556 (74.4) | 1,980 (64.7) | 2,011 (66.5) | 1,942 (64.5) | 3,953 (65.5) |
| <u>Hospital Type</u> |  |  |  |  |  |  |
| None | 46 (4.8) | 77 (3.7) | 123 (4.0) | 126 (4.2) | 121 (4.0) | 247 (4.1) |
| LNU | 540 (55.8) | 1,153 (55.1) | 1,693 (55.3) | 1,693 (56.0) | 1,662 (55.2) | 3,356 (55.6) |
| SCU | 60 (6.2) | 145 (6.9) | 205 (6.7) | 208 (6.9) | 197 (6.5) | 405 (6.7) |
| NICU | 321 (33.2) | 717 (34.3) | 1,038 (33.9) | 997 (33.0) | 1,031 (34.2) | 2,028 (33.6) |
| <u>PPROM</u> |  |  |  |  |  |  |
| No | 775 (80.1) | 1,245 (59.5) | 2,020 (66.0) | 1,960 (64.8) | 1,977 (65.6) | 3,937 (65.2) |
| Yes | 192 (19.9) | 847 (40.5) | 1,039 (34.0) | 1,065 (35.2) | 1,035 (34.4) | 2,099 (34.8) |
| <u>Complications<sup>2</sup></u> |  |  |  |  |  |  |
| No | 736 (76.1) | 1,523 (72.8) | 2,259 (73.8) | 2,244 (74.2) | 2,232 (74.1) | 4,476 (74.1) |
| Yes | 231 (23.9) | 569 (27.2) | 800 (26.2) | 780 (25.8) | 780 (25.9) | 1,560 (25.9) |

Data are n (%).
BMI, body mass index; LNU, local neonatal unit; NND, neonatal death; NICU, neonatal intensive care unit; PPRM, preterm prelabour rupture of membranes; SB, stillbirth; SCU, special care unit; SMID, Scottish Index of Multiple Deprivation.
<sup>1</sup> Extremely preterm was defined as 24<sup>+0</sup> to 27<sup>+6</sup> gestational weeks; very preterm as 28<sup>+0</sup> to 31<sup>+6</sup> gestational weeks; and moderately preterm as 32<sup>+0</sup>-36<sup>+6</sup> gestational weeks.
<sup>2</sup> Complications, defined as the presence at least one of the following conditions: pre-existing or gestational hypertension, pre-eclampsia, pre-existing or gestational diabetes, liver disorders, large-for-gestational age fetus, fetal abnormalities, intrauterine growth restriction, infection, chorioamnionitis, placenta accreta, placenta praevia, placental abruption, antepartum haemorrhage, or rupture of the uterus
<sup>3</sup> The sample sizes listed under the inverse probability weighted birth mode groups and total are the weighted sample sizes.

The characteristics of women who underwent CB or VBB are shown in Table 1. Age, socioeconomic deprivation quintile, hospital facilities, prior pregnancy outcomes, and marital status were similarly distributed across modes of birth. Ethnicity and BMI were similar but there were differences in the proportion of women with missing data. Minor imbalance existed in smoking history at antenatal booking, with a slightly higher proportion of CBs corresponding to non-smokers (58.6%), relative to those in the VBB group (50.1%). There was more pronounced imbalance in parity, as a larger proportion of CBs were first births (57.4%) than in the VBB group (43.5%), and PPROM, which affected 40.5% of women in the CB group compared to 19.9% in the VBB group. Large imbalance in the distribution of prematurity existed between the two birth groups, with a much larger proportion of extremely preterm births (gestation 24^+0^-27^+6^ weeks) in the VBB group (36.2%) relative to the CB group (7.2%) and a smaller proportion of moderately preterm births (gestation 32^+0^-36^+6^ weeks) in the vaginal group (43.8%) compared to the CB group (74.4%). While some specific complications such as pre-eclampsia were imbalanced across birth groups (Supplemental Table 6), the presence of any complication, a composite measure of all complications including pre-eclampsia, was similarly distributed across mode of birth groups.

Following inverse probability of treatment weighting, sufficient balance in measured confounders was attained between birth modes, based on standardised mean difference values (Figure 3). Imbalances in booking smoking history, parity, and gestation were alleviated with the implementation of inverse probability weighting (Table 1). The total effective estimated sample size, i.e., the estimated sample size needed for a real-life randomised controlled trial to have the same power as the present emulated trial analysis, was 1,498 women across both birth modes.

**Figure 3.**
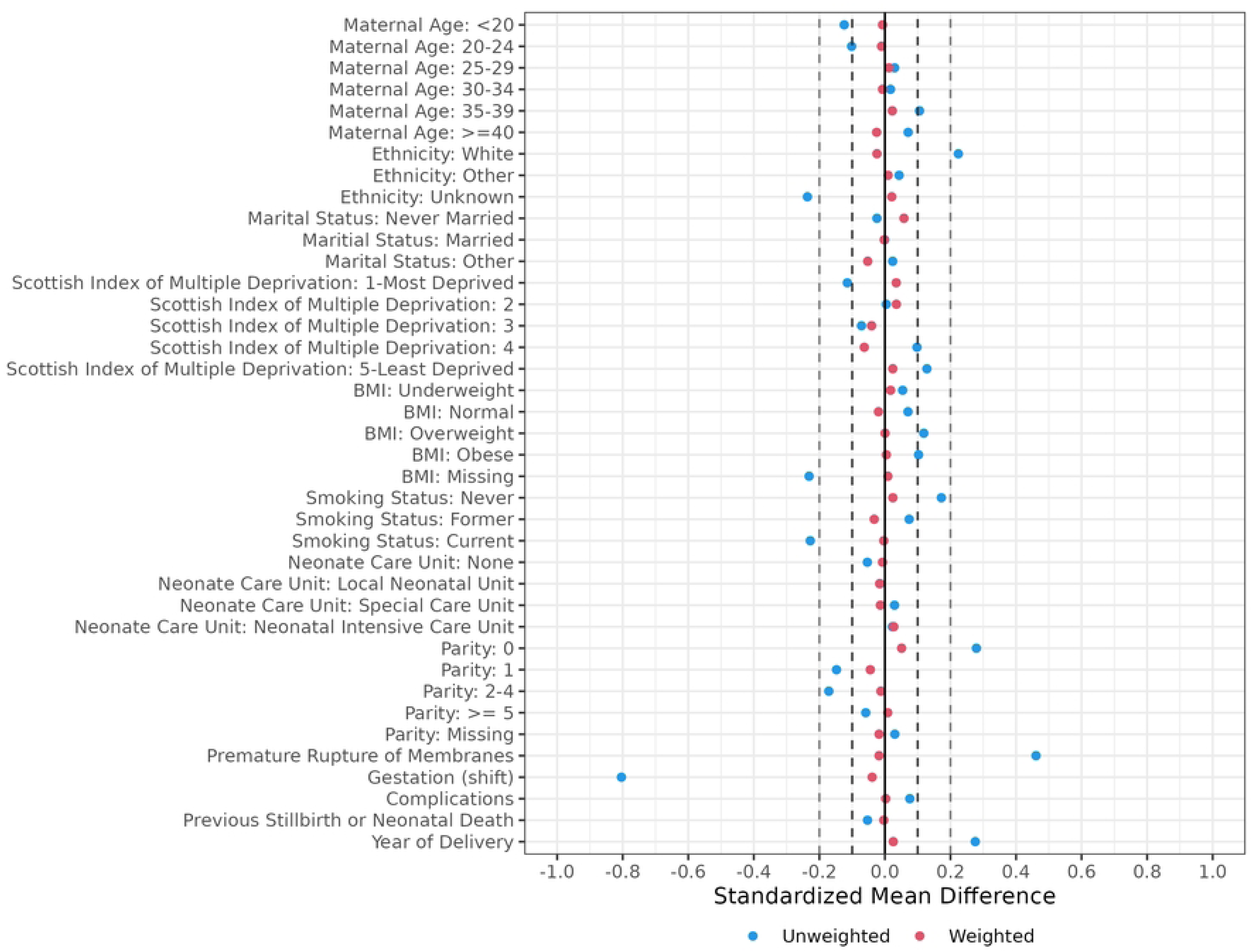
Standardised mean differences before and after inverse probability weighting. This figure demonstrates the standardised mean differences between patients who were treated with CB or no CB for all covariates in the unweighted study population (blue dots) and after applying the weights derived from inverse probability weighting (red dots). The dark dashed lines denote standardised mean differences between-0.1 and 0.1, and the light dashed lines denote standardised mean differences between-0.2 and 0.2. In the weighted sample, none of the covariates exceeded 0.1, indicating sufficient balance in baseline characteristics between women who underwent CB or had a VBB. (BMI, body mass index, in kg/m^2^).

Overall, 92.2% neonates survived the first 28 days of life, with approximately 1.3% of deliveries resulting in intrapartum stillbirths, 5.0% resulting in early neonatal death (death within 7 days of birth), and 1.5% resulting in late neonatal death (death between 7 to 28 days of life). Extended perinatal death occurred amongst 2.6% of CBs and 18.9% of VBBs (Table 2).

**Table 2.** Birth outcomes, by mode of birth.

| **Outcome** | **Vaginal Birth**<br>N = 967 | **Caesarean Birth**<br>N = 2,092 | **Total**<br>N = 3,059 |
| --- | --- | --- | --- |
|  | n (%) | n (%) | n (%) |
| Extended perinatal death | 183 (18.9) | 55 (2.6) | 238 (7.8) |
| Stillbirth | 34 (3.5) | 5 (0.2) | 39 (1.3) |
| Early neonatal death (0-6 days) | 117 (12.1) | 36 (1.7) | 153 (5.0) |
| Late neonatal death (7-28 days) | 32 (3.3) | 14 (0.7) | 46 (1.5) |

519 Data are n (%).

Babies delivered by CB had lower odds of perinatal death by 68.8% (OR 0.31, 95% CI: 0.25 to 0.39). While the protective impact of CB remained significant across all gestations, the effect of CB on extended perinatal mortality was modified by gestational age at birth. The magnitude of protective effect of CB on extended perinatal mortality increased by 9% for each additional week gestation (OR 1.09; 95% CI 1.02 to 1.18 (Figure 4, Table 3, supplemental Table 7). CB at 24 weeks gestation decreased the odds of perinatal death by 47.7% (OR: 0.53, 95% CI: 0.35 to 0.78). At 36 weeks’ gestation, the impact of CB was largest, decreasing the odds of perinatal death by 82.1% (OR: 0.18, 95% CI: 0.10 to 0.32).

**Figure 4.**
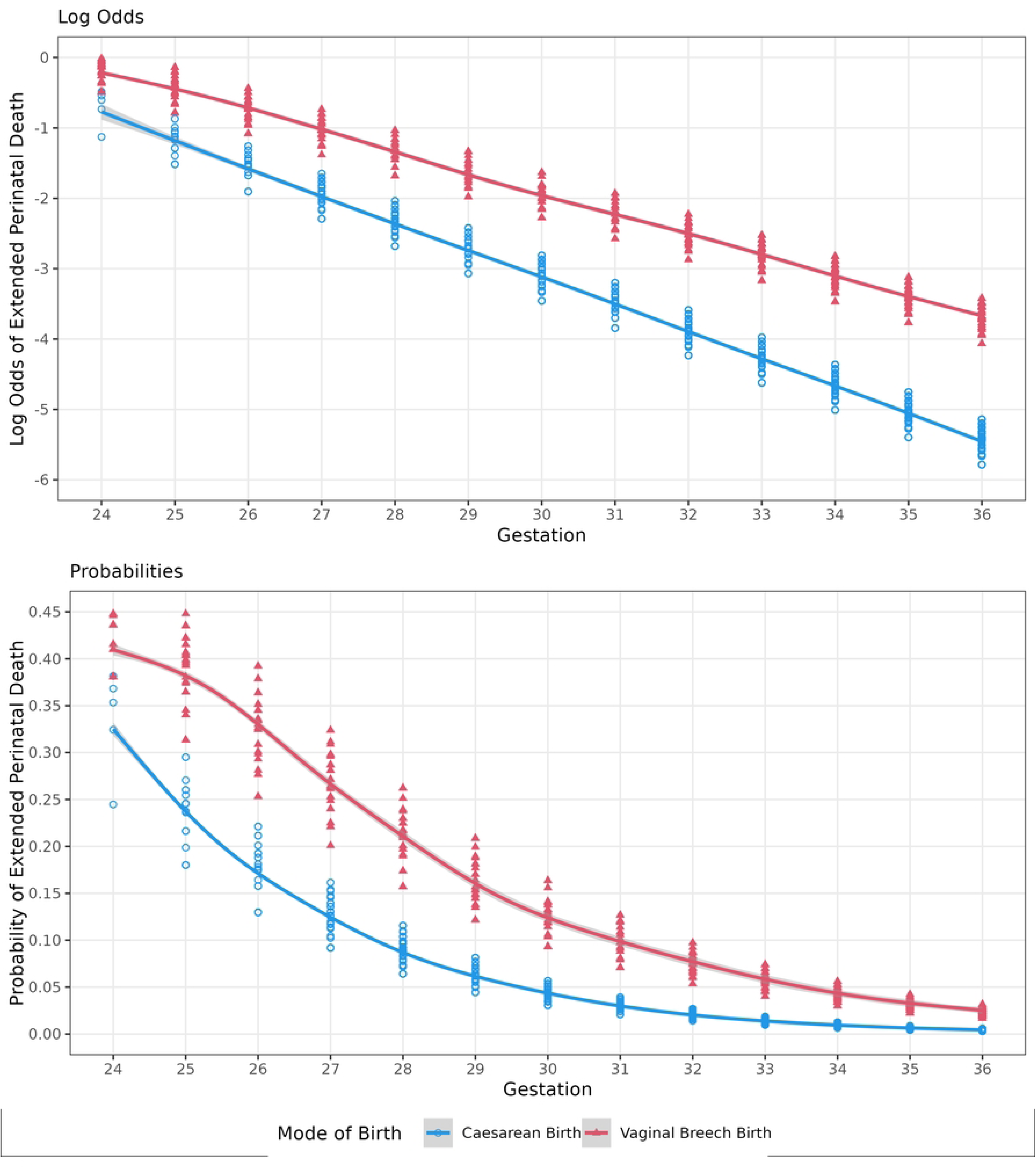
Log odds and probability of extended perinatal mortality, by mode of birth. This figure demonstrates the change in estimated causal impact of mode of birth on extended perinatal mortality and the likelihood of perinatal mortality across different gestations. The log odds highlight the increasing impact of CB on perinatal survival as gestation increases, while the probabilities emphasise the change in diminishing likelihood of perinatal death as gestational age increases.

**Table 3.**
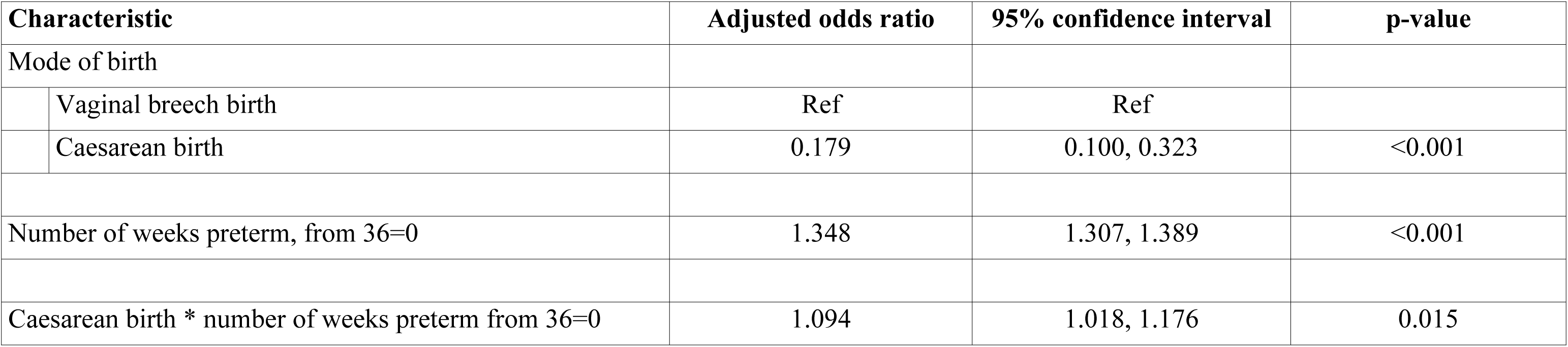
Model Results.

This relationship was complicated by the change in probability of extended perinatal death across gestational age (Figure 4, Table 3, supplemental Table 7). The odds of perinatal death increased by 1.34 (95% CI 1.31, 1.39) with each additional week of prematurity. Whilst the magnitude of the decrease in the odds of perinatal death in women undergoing CB was larger at more advanced gestational age, the odds of perinatal death were much lower closer to term compared to the extreme preterm period. This is reflected in the number needed to treat (NNT) estimates, where among women delivering at 36 weeks’ gestation, 88 would need to undergo CB to prevent one extended perinatal death, while among women delivering at 24 weeks gestation only seven would need to undergo a CB to prevent one extended perinatal death (see Supplemental Table 7 for NNT estimates for each gestational week).

The array approach sensitivity analysis to evaluate impact of unmeasured confounders demonstrated that the analysis is robust to unmeasured confounders in most cases. Considering all preterm breech births (24 to 36 weeks’ gestation), in order to reverse our results, an unmeasured factor would need to meet the following extreme conditions: it is associated with a ∼6-fold increase in the odds of perinatal mortality, have a prevalence of ≥75% in the VBB group, and have a concurrent prevalence of ≤20% in CB group (Figure 5). At 24 weeks, the unmeasured factor would need to be associated with a ∼3-fold increase in the odds of perinatal mortality, have a prevalence of ≥50% in the VBB group, and have a concurrent prevalence of ≤20% in the CB group.

**Figure 5.**
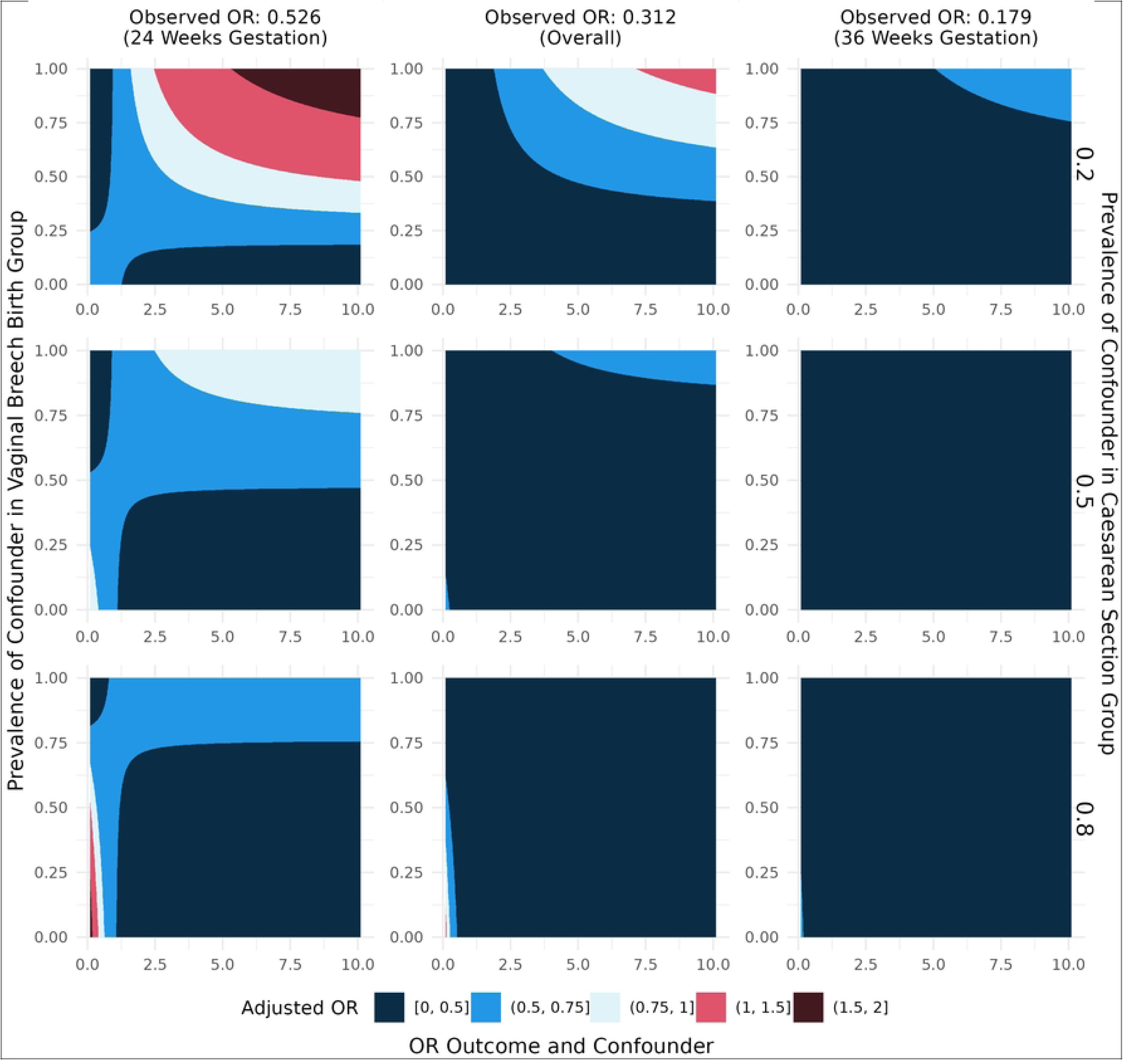
Sensitivity analysis array approach. This figure illustrates the impact of an unmeasured confounding factor on the final analysis. The magnitude of the estimated impact of CB on extended perinatal mortality is depicted by the colour of the plots, with red hues denoting a reverse in the analysis results. Within each plot, the vertical axis varies the prevalence of the confounding variable in the vaginal breech birth group, while the rows of the graphs in descending order increases the prevalence of the confounding variable in the CB group. Within each plot, the horizontal axis varies the strength of the relationship between the confounding variable and extended perinatal mortality (<1 implies a protective effect while >1 implies a risk factor for extended perinatal mortality). The columns from left to right offer establish the estimated effects of CB on extended perinatal mortality at 24 weeks’ gestation, from 24-36 weeks’ gestation, and at 36 weeks’ gestation (OR, odds ratio).

## Discussion

In target trial emulation using Scottish observational data and deploying a per-protocol analysis (by actual mode of birth), CB was associated with a significant reduction in perinatal mortality compared to vaginal birth amongst women presenting in spontaneous-onset labour at 24-36 gestational weeks with a singleton live baby presenting breech. Whilst CB was associated with reduced perinatal mortality throughout the entire preterm period, the magnitude of this protective effect increased, whilst the risk of perinatal death decreased, with increasing gestational age. To our knowledge, our analysis is the first to apply trial emulation methodology to explore the effect of CB on perinatal mortality for preterm breech labour.

The study has several strengths. It applies causal inference methods to emulate a clinical trial by applying the principles of clinical trial design and adjusting for a range of measured confounders to limit the risk of immortal time bias and confounding by indication. Our target trial specifically considered a subset of women who attend in spontaneous-onset (unplanned) labour. In this scenario clinicians and parents are required to decide on mode of birth within a relatively short period time, which may limit indication bias and reflects a real-life clinical challenge for the practicing obstetrician, especially at earlier gestation. Furthermore, the trial was emulated using comprehensive observational data spanning across 22 years and adjusting for year of birth, clustering by hospital, and levels of neonatal care. In the absence of randomised trial data, trial emulation may be the most robust and feasible approach to explore the effects of CB on perinatal mortality, thereby enabling enhanced patient-clinician decision-making and providing incentive to other researchers to evaluate impacts of CB on perinatal outcomes by applying trial emulation methodology to observational data.

There are several limitations to this study. Firstly, our observational data set limited us to a per protocol analysis, wherein we emulated the trial by actual mode of birth, i.e., the treatment the trial participants ultimately received. This approach carries inherent selection bias,^8^ and does not reflect those cases were women assigned to VBB ultimately undergo CB for, e.g., concerns over fetal distress in labour, and conversely, women assigned to CB may proceed to a VBB before CB can be organised. Second, we were unable to disaggregate footling breech births. Third, we conducted a complete case analysis, i.e., only including participants with complete data, which could incur bias. Furthermore, our trial was emulated using data from Scotland, limiting generalisability of our findings to other settings where skill sets to support VBB and availability of CB may differ. Moreover, we were unable to adjust for palliative treatment, which some parents may have chosen at early gestation, and the occurrence of which could have been more frequent amongst women who birthed vaginally. Lastly, we are uncertain of the direction of bias that may arise from unknown and unmeasured confounding factors that we were unable to adjust for, although our sensitivity analysis suggests it is unlikely missing confounders would completely reverse our results.

The characteristics of our study population (spontaneous onset of labour +/-PPROM) and methodological approach most closely align with recent analyses of European observational data that used approximates of neonatal mortality as the primary outcome and applied propensity score matched or weighted statistics to adjust for indication bias.^13 17^ Schmidt et al. reported a 50% reduction in the odds of the composite of in-hospital mortality intraventricular haemorrhage and cystic periventricular leukomalacia in women who had a CB (analysis by actual mode of birth) at 24-31 weeks (572 pregnancies without antenatal complications, 11 European countries) compared to VBB when deploying conventional mixed-effects adjusted logistic regression analysis. This association was no longer statistically significant in propensity score matched models, or when analysed by institutional policy (intended mode of birth). Lorthe et al reported no benefit of planned CB over VBB in terms of neonatal survival in propensity score matched analysis (26-34 gestational weeks, 390 births). Other observational studies evaluating the effect of CB on neonatal death produced mixed results but differed substantially in their approach to analysis (actual or intended mode of birth), cohort selection, selection of prognostic factors, and approaches to adjusting for confounders.^11 14 19 44 45^

Our target trial suggests that in a per-protocol analysis CB reduces the risk of extended perinatal mortality for singleton preterm breech from 24 to 36 gestational weeks. This protective effect increases, and the risk of perinatal death decreases, with increasing gestational age. Therefore, fewer CBs are required to prevent a perinatal death for women in extreme preterm breech labour compared to women in late preterm breech labour. Whilst the effect of CB on perinatal mortality estimated from per-protocol analysis tends to be larger than those observed in an intention-to-treat approach, the reduction in perinatal mortality observed in our trial is substantial. Our study considers the most significant adverse infant outcomes, intrapartum stillbirth and neonatal death, but does not consider other infant outcomes and maternal morbidity. This, and the smaller effect sizes observed in intention to treat analyses must be considered in decision-making and discussions with parents.^25^ Furthermore, the findings apply to a sub-set of women in spontaneous preterm breech labour with no previous CB. Effect sizes may vary depending on availability of clinical skills to support CB and VBB in a specific setting,^46^ and findings should not be extrapolated to settings where access to timely and safe CB is limited. Lastly, findings should not be used to promote a particular mode of birth over another but allow patients to make a more informed choice as to how they wish to deliver their preterm breech baby.

We encourage similar trial emulations, especially in settings with a comparable health systems architecture such as in Scotland, preferably using observational data that captured both actual and intended mode of birth reliably. Aligning definitions of preterm breech phenotypes and cohort selection criteria to allow multi-jurisdictional studies of the impacts of CB on health outcomes could greatly assist with deepening our understanding by allowing a degree of comparability. Furthermore, longer term impacts of CB for child and maternal health, including outcomes in subsequent pregnancies are worth exploring further.^47^ A different approach to choosing the optimal mode of birth for women in spontaneous preterm labour may be to determine the effect sizes of CB in specific sub-populations (e.g., nulliparous versus parous women), but stratification requires larger data sets to enable careful study of this relatively rare clinical scenario and outcome. Such analyses may enable more nuanced evidence-based counselling, considering a range of clinical characteristics, and permitting a more case-specific approach.^10^

## Data Availability

Data can be obtained through an application made to the Public Health Scotland electronic Data Research and Innovation Service (PHS-eDRIS) of Public Health Scotland (https://www.informationgovernance.scot.nhs.uk/pbpphsc/home/for-applicants/)

## Acknowledgements

We thank the eDRIS Team at Public Health Scotland for their support with obtaining approvals, provisioning and linking data, and the use of the secure analytical platform within the National Safe Haven.

## Contributors

HWU and SB conceived the study and obtained research funding. RA, HWU, SB, AAL, BS, CM, SJS and SAU contributed to the design and conduct of the study. RA, CM, BS, AAL, CC, KKK, AFF, SB, HWU and SJS contributed to the analysis of data. All authors contributed to the interpretation of the data and presentation, review, critical revision and approval of the manuscript. RA and HWU are guarantors. The corresponding authors attests that all listed authors meet authorship criteria and that no others meeting the criteria have been omitted.

## Funding

HWU was supported by a Glasgow Children’s Hospital Charity Research Fund Project Support Grant (GCHC/PSG/2018/10) and a Menzies School of Health Research Maple-Brown Foundation Future Leaders Fellowship. SB was supported by a National Health Service (NHS) Fife Research and Development Bursary. SJS was funded by a Wellcome Clinical Career Development Fellowship 209560/Z/17/Z. SAU was funded by a National Health Service (NHS) Scotland Research (NRS) Fellowship. The funders had no role in considering the study design or in the collection, analysis, and interpretation of data, the writing of the report, or the decision to submit the article for publication.

## Competing interests

All authors have completed the ICMJE uniform disclosure form at www.icmje.org/disclosure-of-interest/. SJS declares she is the Program Director of the Wellcome Leap In Utero Program. She has received grant funding (paid to her institution) from Wellcome, MRC, CSO Scotland and Tommy’s. She has received consultancy fees from Natera and Norgine and honoraria for educational lectures from Hologic. All other authors declare no support from any organisation for the submitted work other than that described above; no financial relationships with any organisations that might have an interest in the submitted work in the previous three years; no other relationships or activities that could appear to have influenced the submitted work.

## Notes

### Funding Statement

Yes

### Author Declarations

Ethical approval for usage of anonymised routinely collected data to enable this trial emulation was granted by the National Health Service Scotland Public Benefit and Privacy Panel for Health and Social Care (1819-0119) and the University of St Andrews Teaching and Research Ethics Committee (MD15778). Ethical approval included a waiver of informed written consent for individuals included within the study. Access to the datasets by the authorised researchers was exclusively within a Safe Haven environment after deidentification of the records.

